# Selective Removal of Endometriotic Lesions Using CUSA Clarity in Ovarian Endometriomas: A Case-Based Histopathological Study

**DOI:** 10.1101/2025.10.12.25334539

**Authors:** Tatsuhito Kanda, Takashi Hosono, Ai Sato, Yukari Maeda, Iwao Yasoshima, Yuya Sakai, Haruki Kasama, Kayo Kayahashi, Kyosuke Kagami, Takashi Iizuka, Kaoru Abiko

## Abstract

**Objective:** To evaluate the feasibility of selective removal of endometriotic lesions using the Cavitron Ultrasonic Surgical Aspirator (CUSA® Clarity) in ovarian endometriomas, with a focus on histological preservation of normal ovarian tissue.

**Methods:** We analyzed tissue from a woman in her early 30s who underwent laparoscopic surgery for an ovarian endometrioma measuring approximately 7 cm after preoperative dienogest therapy. Resected cyst wall specimens were divided into five parts, each assigned to a Tissue Select® setting (0–4). Samples were scraped with CUSA, followed by histological and immunohistochemical evaluation (H&E, Sirius Red, CK7, CD10).

**Results:** Endometriotic lesions (epithelial and stromal cells) were effectively removed across all settings. At higher Tissue Select settings (3–4), preservation of surrounding tissue was superior, with minimal vacuolization compared to lower settings (0–2). Primordial follicles were observed approximately 600 μm beneath the surface, highlighting the importance of limiting cavitation depth.

**Conclusion:** CUSA Clarity enabled selective removal of endometriotic lesions with relative preservation of normal ovarian tissue, particularly at higher Tissue Select settings. This novel approach may represent a fertility-preserving alternative to cystectomy or laser ablation in the management of ovarian endometriomas. Further studies are warranted.

## Introduction

Surgical treatment is often considered for ovarian endometriomas in cases complicating infertility to improve fertility (1). In ovarian endometriomas, cystectomy can inadvertently excise the normal ovarian tissue, causing bleeding, local hypoxia, and thermal injury from hemostatic energy devices, all of which may impair ovarian function (2). Recently, laser ablation has been reported to better preserve ovarian function than cystectomy, as indicated by higher postoperative antral follicle counts and anti-Müllerian hormone (AMH) levels (3). Although its efficacy on pregnancy and recurrence rates remain inconclusive, fertility-preserving surgical approaches are increasingly sought.

Cavitron Ultrasonic Surgical Aspirator (CUSA®) is a surgical device that uses ultrasonic cavitation to selectively fragment and aspirate soft tissue. It preferentially targets water-rich tissues such as fat while sparing collagen-rich structures. We hypothesized that the CUSA system, particularly the Clarity model equipped with a five-step Tissue Select® mode, could selectively remove endometriotic lesions while minimizing damage to normal ovarian tissue. To date, only one report has described its use for ovarian endometriomas, without histological evaluation (4). We histologically evaluated the selective effect of CUSA Clarity on endometriotic and ovarian tissue in surgical specimens.

## Methods

This exploratory study used tissue obtained from a single surgical case (Fig. 1). A woman in her early 30s underwent laparoscopic cystectomy for an ovarian endometrioma approximately 7 cm in diameter, following one month of preoperative hormone therapy with dienogest. The cyst wall was primarily resected using forceps and scissors, with minimal energy device use. After confirming the absence of solid components, part of the resected cyst wall was sampled for analysis. The specimen was divided into five parts, each assigned to a Tissue Select setting (0–4), a mode available in the CUSA Clarity system. The central region of each sample was gently scraped using CUSA (Supplemental Video). The samples were fixed in formalin and paraffin embedded. H&E staining, Sirius Red staining, and immunohistochemistry (CK7, CD10) were performed. Histological assessment was performed to evaluate the removal of endometriotic lesions and the preservation of normal ovarian tissue. This study was approved by the Clinical Research Ethics Committee of Kanazawa University (approval number: 114898-1), and a written informed consent was obtained.

**Figure 1.**
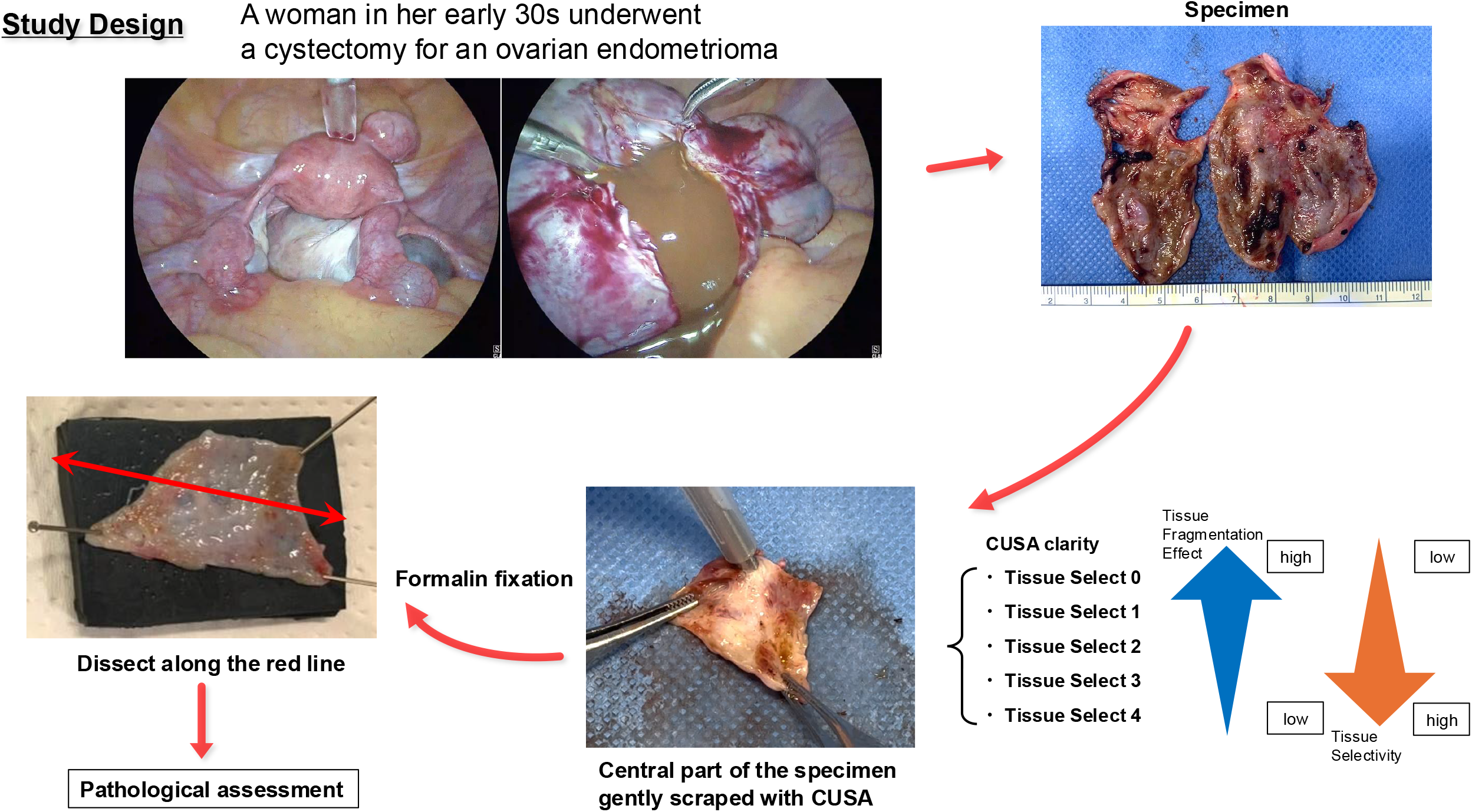
Overview of study design. Tissue from a laparoscopically resected ovarian endometrioma was divided into five parts and assigned to different Tissue Select settings (0–4) of the CUSA Clarity system. The central region of each specimen was gently scraped with CUSA and processed for histological analysis, including H&E, Sirius Red, and immunohistochemical staining (CK7 and CD10).

## Results

Immunohistochemistry confirmed epithelial (CK7), stromal cells (CD10) of endometriosis, and a dense collagenous layer (Sirius Red) beneath the lesions (Supplemental Fig. 1). CUSA, using the Tissue Select setting 4, effectively removed the epithelial and stromal cells of endometriosis from the scraped area without tissue damage (Fig. 2). In all Tissue Select settings, endometrioid lesions positive for CD7 and CD10 were effectively removed from the scraped areas (Supplemental Fig. 2). Residual lesions were observed at the scraped margins, confirming the presence of disease at the non-scraped side of the margin in each sample (Supplemental Fig. 3).

**Figure 2.**
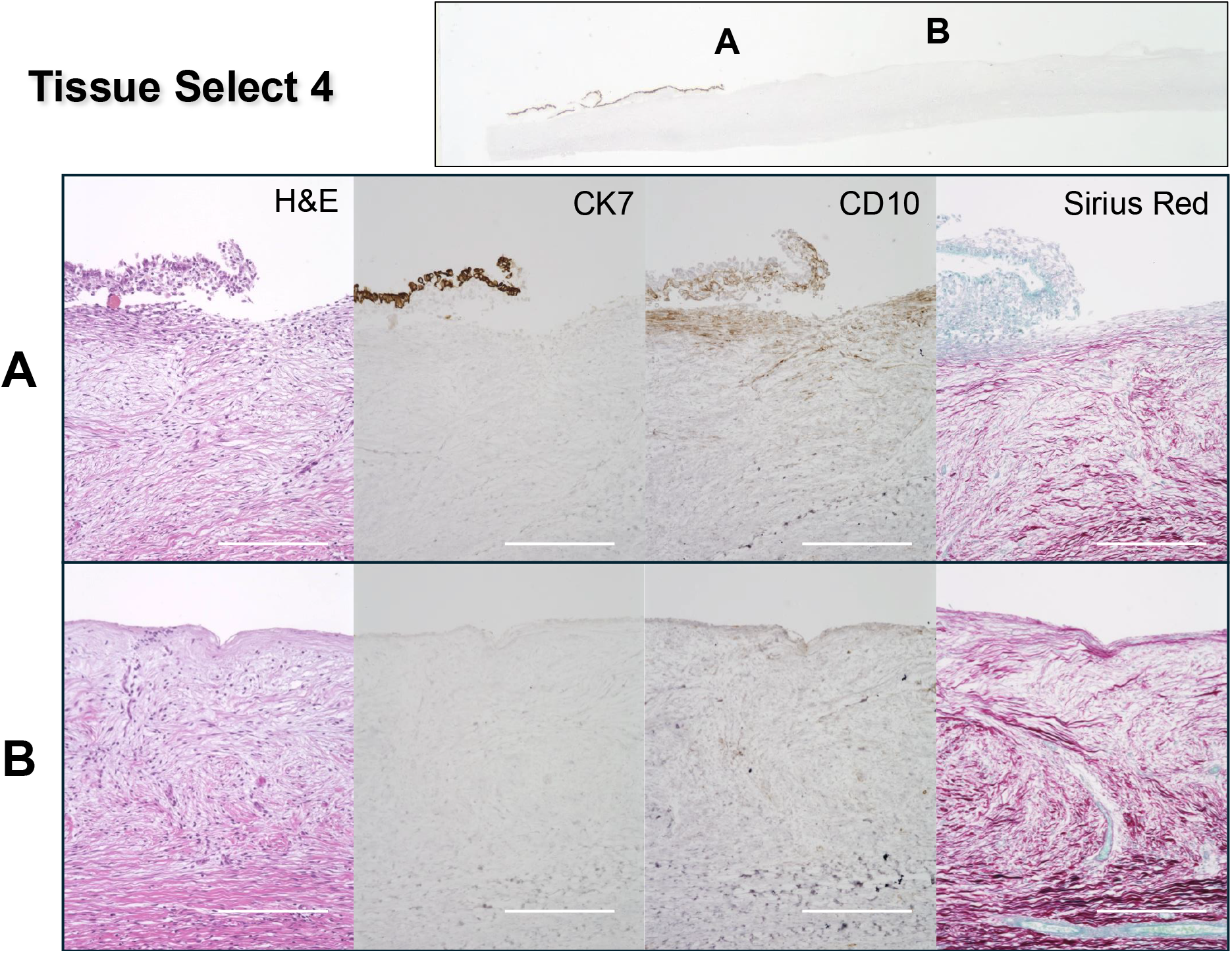
Histological findings at Tissue Select setting 4. (A) Scraped margin showing residual endometriotic tissue on the unscraped side (to the left) with preserved staining quality. (B) Central area of the scraped region showing minimal tissue damage and effective removal of the endometriotic lesion.69 Main images were taken at ×200 magnification. Scale bar = 200Lμm. The upper right image shows CK7 immunostaining at ×10 magnification.

Damage to normal ovarian tissue varied by Tissue Select setting (Supplemental Fig. 4). At setting 0, vacuolization and tissue degeneration extended to depths of 200–400 μm. Similar effects were observed at settings 1–2, generally within 200 μm. At settings 3–4, effects were more limited.

In some samples, primordial follicles were identified approximately 600 μm from the surface, underscoring the importance of limiting cavitation and thermal effects to superficial layers (Supplemental Fig. 5).

## Discussion

This exploratory study provides preliminary histopathological evidence that the Cavitron Ultrasonic Surgical Aspirator (CUSA® Clarity) may enable selective removal of endometriotic lesions in ovarian endometriomas while preserving adjacent ovarian tissue. At higher Tissue Select settings (3–4), epithelial and stromal components of endometriosis were effectively removed, whereas damage to surrounding collagen-rich stroma was limited. Importantly, primordial follicles were identified at depths as shallow as 600 μm beneath the cyst wall, underscoring the clinical relevance of minimizing tissue injury during fertility-preserving surgery.

Conventional cystectomy has long been considered the standard surgical approach for ovarian endometriomas but carries the inherent risk of excising normal ovarian tissue together with the cyst wall. This can lead to decreased ovarian reserve, as reflected by reductions in antral follicle counts and serum AMH levels postoperatively (5, 6). Energy-based hemostasis may further exacerbate cortical injury through thermal effects. In contrast, ablative techniques such as COL laser or plasma energy vaporization have been reported to better preserve ovarian function, though concerns remain regarding recurrence rates and long-term fertility outcomes (3, 7).

Recent studies evaluating the depth of tissue ablation with alternative energy sources provide useful benchmarks for fertility-preserving surgery. In a pilot study of argon plasma coagulation for ovarian endometriomas (APC-ENDO), the mean depth of ablation was reported to be about 0.8 mm, suggesting that laser- and plasma-based modalities can achieve controlled ablation to a limited depth within the cyst wall (8). Similarly, a prospective multicenter trial investigating diode laser vaporization of ovarian endometriomas (OMAlaser) demonstrated ablation depths of around 500 *μ*m. Importantly, this study also reported favorable outcomes in terms of ovarian reserve recovery, fertility preservation, and low recurrence rates, even in patients with advanced endometriosis (9).

Histologic analysis has shown that the penetration of endometriosis into cyst walls is highly variable, with a mean of approximately 0.6mm ± 0.4mm and 99% of cases measuring within 1.5 mm, but occasionally reaching up to 2.0 mm (10). This variability has important surgical implications. In cysts with relatively thick walls, laser vaporization may not penetrate deeply enough to eradicate endometriotic tissue completely, resulting in residual disease. Conversely, when the cyst wall is thin, laser ablation may extend too deeply and risk excessive thermal injury to the ovarian cortex. By contrast, the mechanism of CUSA—selective disruption of water-rich endometriotic tissue with preservation of collagen-rich fibrotic structures—may provide an intrinsic safety advantage. Cavitation tends to halt at fibrotic boundaries, thereby reducing the likelihood of over-treatment and minimizing collateral damage to adjacent healthy ovarian tissue. In the current study, our pathological examination demonstrated that CUSA completely removed endometriotic lesions while preserving the ovarian parenchyma—composed of ovarian fibroblasts covering primordial follicles—without damage.

This study has several limitations. The analysis was based on a single surgical specimen, without correlation to postoperative ovarian reserve or fertility outcomes. The selective effect was demonstrated only ex vivo, and the in vivo impact on hemostasis, bleeding risk, and recurrence remains unknown. Moreover, optimal CUSA settings for balancing lesion removal with tissue preservation require further validation.

In conclusion, this proof-of-concept study indicates that CUSA Clarity, particularly at higher Tissue Select settings, has the potential to achieve fertility-preserving removal of endometriotic lesions in ovarian endometriomas. Further prospective studies with larger patient cohorts and clinical outcome measures are warranted to establish its safety and efficacy compared with established surgical modalities.

## Data Availability

All data generated or analyzed during this study are included in this published article. Additional information is available from the corresponding author on reasonable request.

## Ethics

This study was approved by the Clinical Ethics Committee of Kanazawa University (IRB approval number 114898-1). Written informed consent was obtained from the patient.

