## Supplementary figures and images for "Selective Removal of Endometriotic Lesions Using CUSA Clarity in Ovarian Endometriomas: A Case-Based Histopathological Study"

### Supplemental Figure 1

#### Tissue Select 4

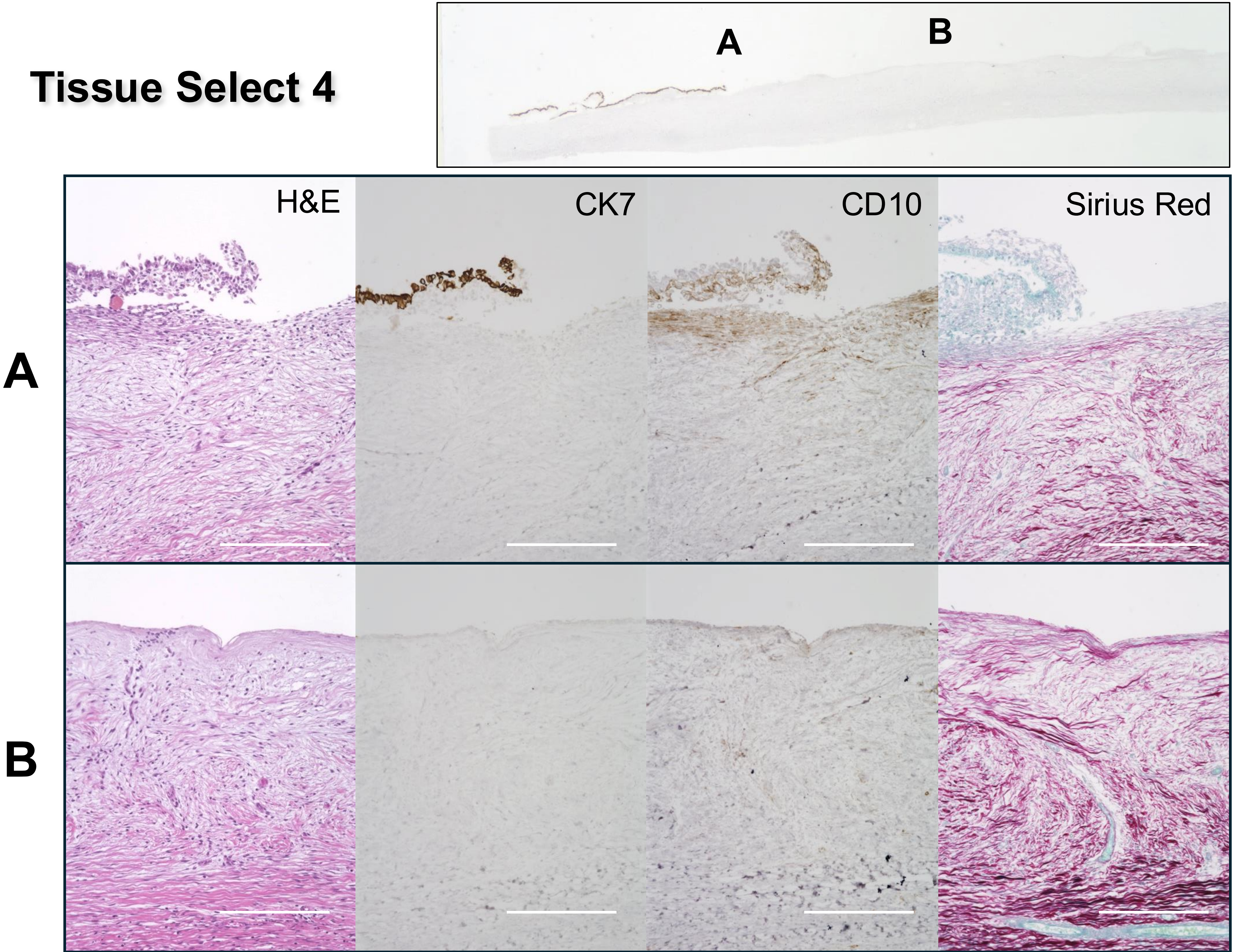

### Supplemental Figure 2

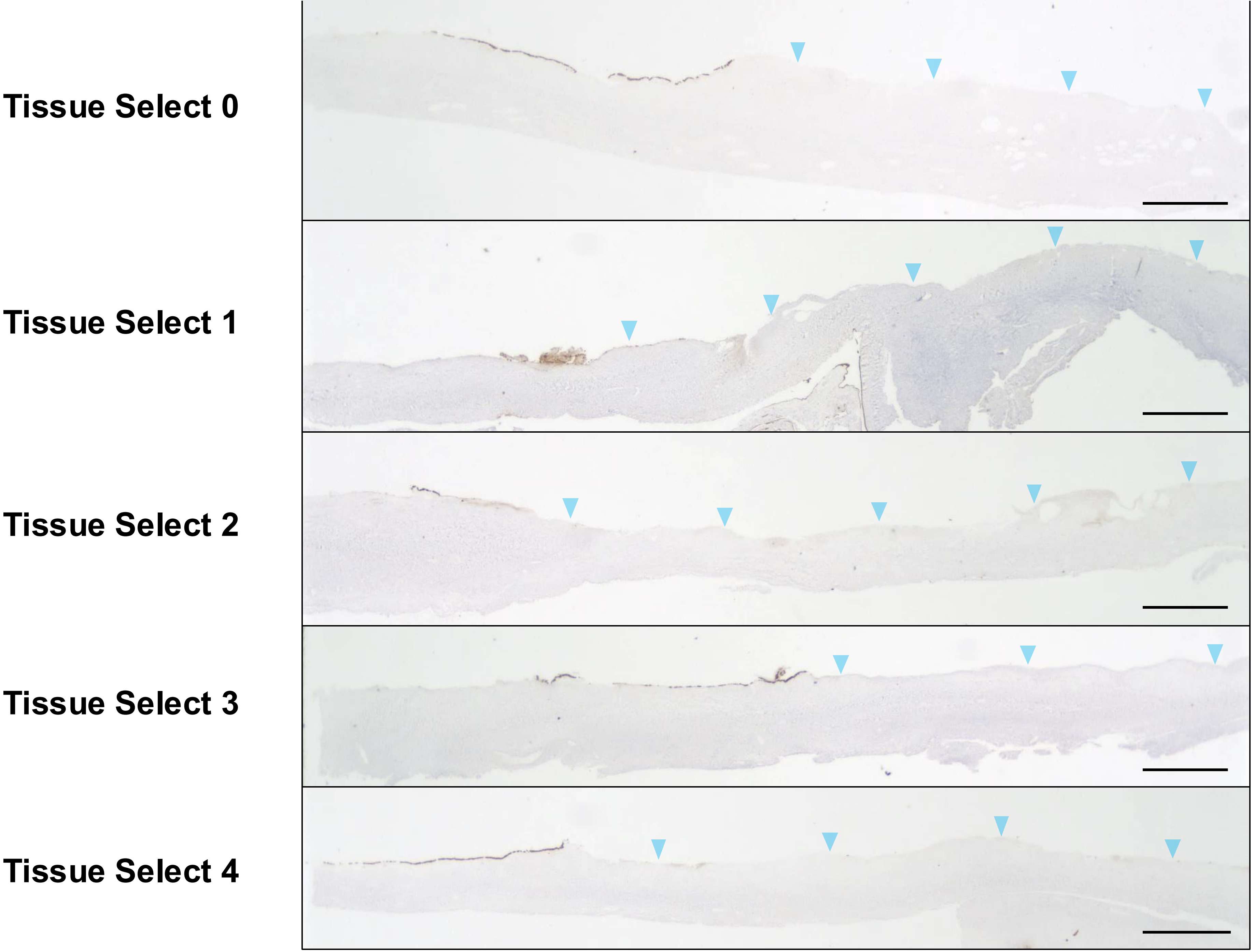

### Supplemental Figure 3

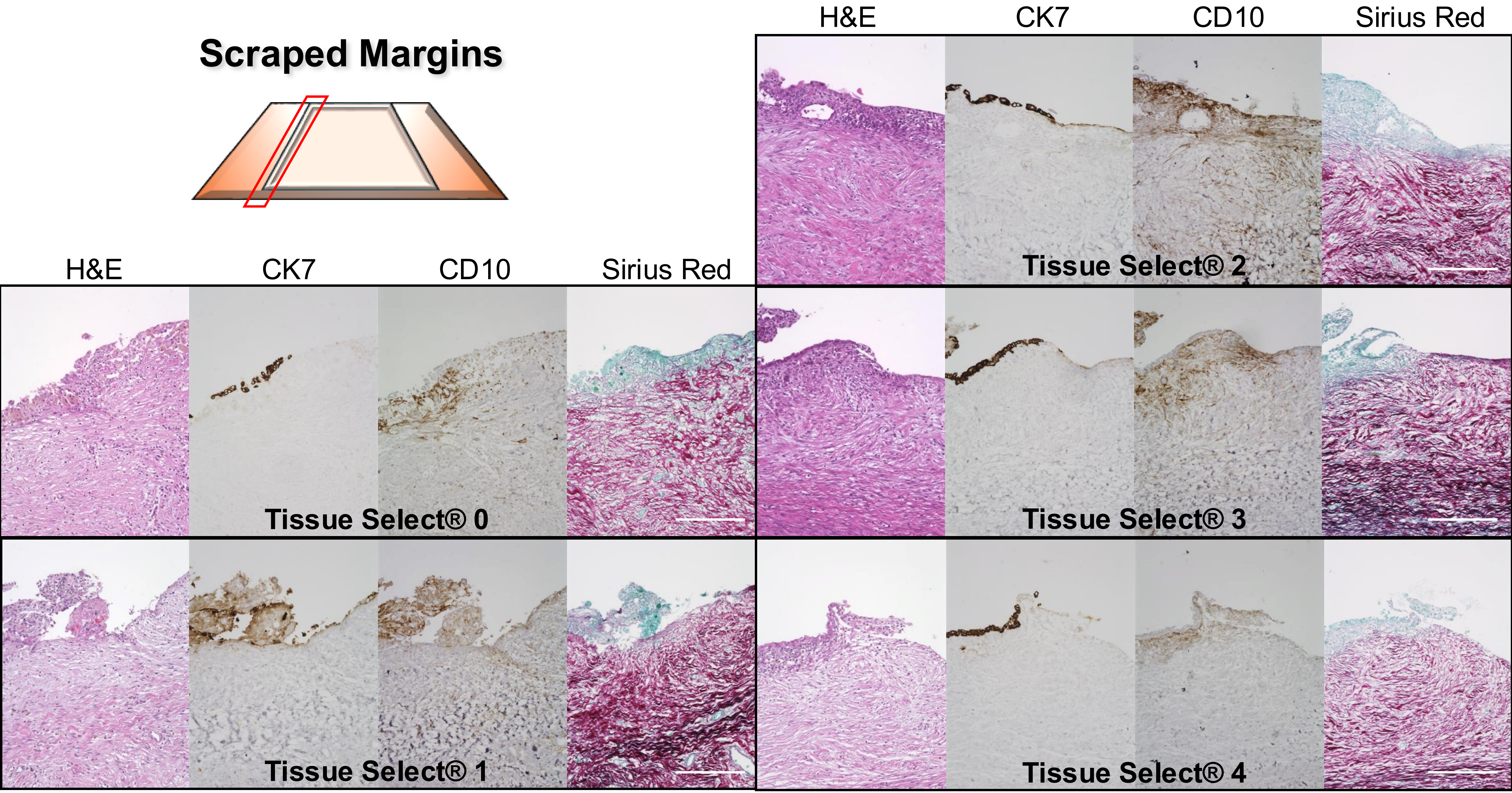

### Supplemental Figure 4

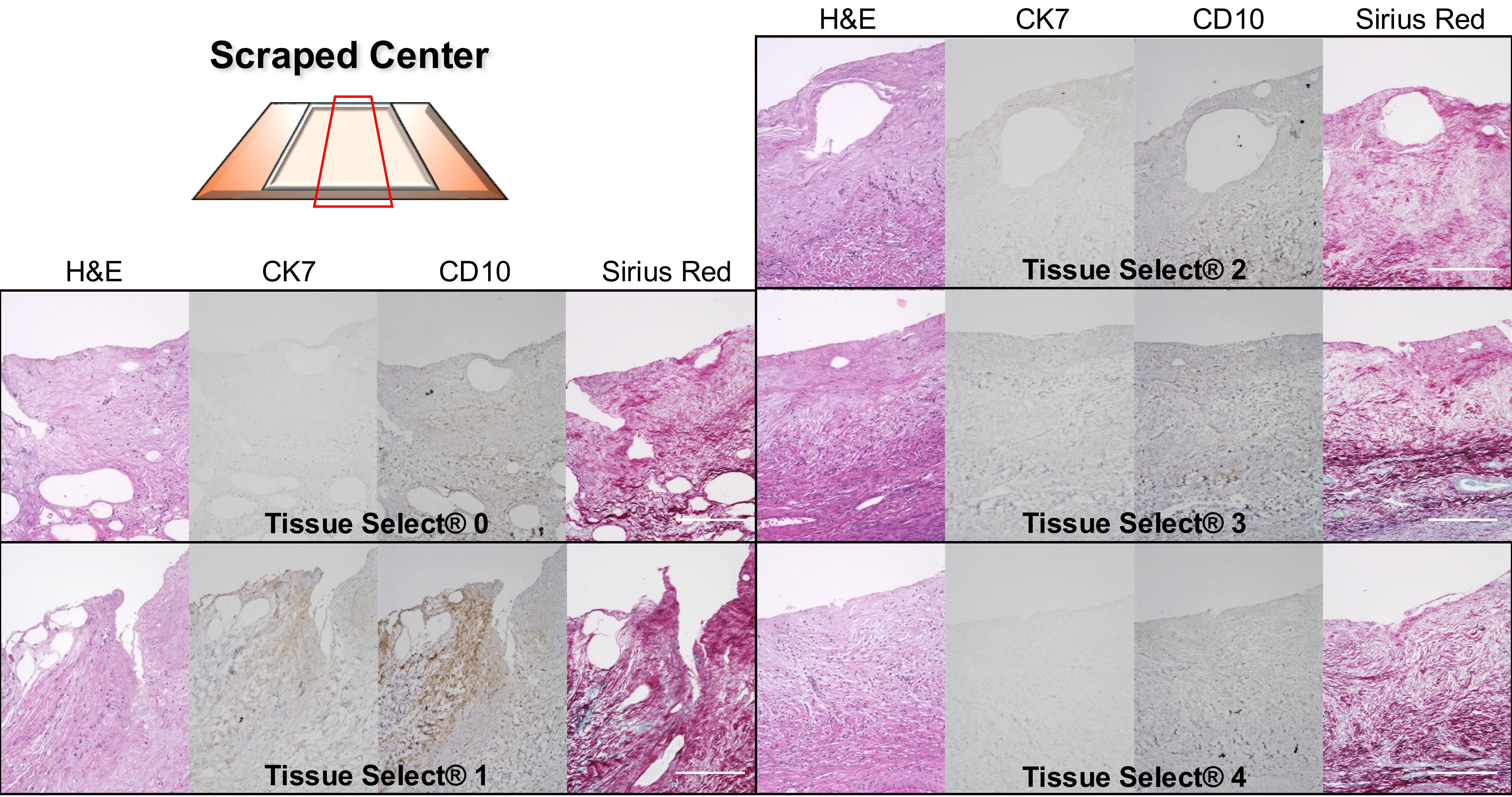

### Supplemental Figure 5

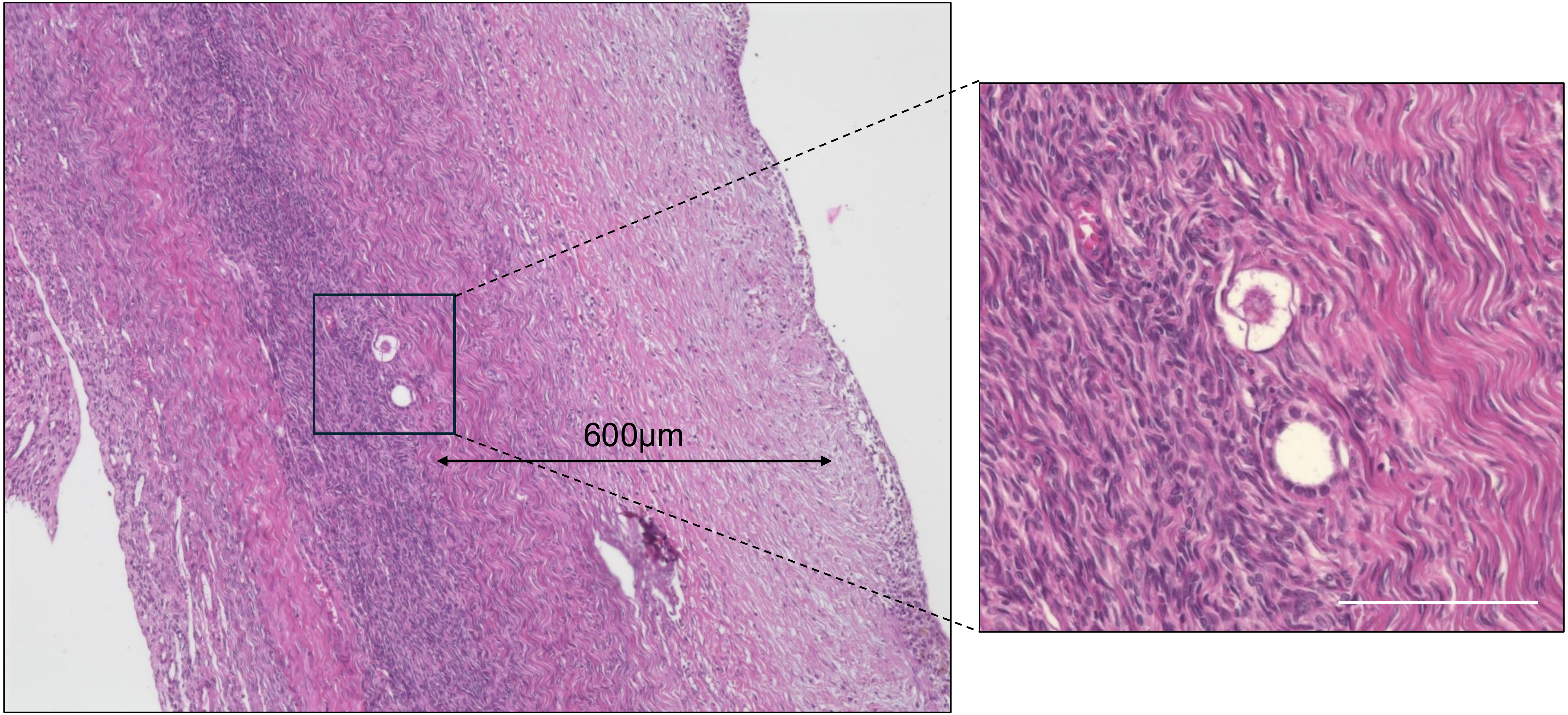
