## Supplementary_Figure&Video_Legends for "Selective Removal of Endometriotic Lesions Using CUSA Clarity in Ovarian Endometriomas: A Case-Based Histopathological Study"

**Supplementary Figure Legends**

**Supplemental Figure 1. Histological features of an endometriotic lesion in the control area.**
Representative images at ×200 magnification show CK7- and CD10-positive staining in epithelial and stromal cells of endometriosis. Sirius Red staining reveals a pale blue to light green lesion overlying a dense, red-stained collagen-rich fibrotic layer. Scale bar = 200 μm.

**Supplemental Figure 2. CK7 immunostaining of scraped areas at Tissue Select settings 0–4.**
Representative images at ×10 magnification illustrate the absence of CK7-positive epithelial cells of endometriosis in the scraped regions in all settings. Blue arrowheads indicate the areas scraped with CUSA. Scale bar = 1000 μm.

**Supplemental Figure 3. Scraped margins at Tissue Select® settings 0–4.**
Representative images of the scraped margins show residual endometriotic lesions in all settings, confirming the presence of disease at the non-scraped side of the margin in each sample. All images were taken at ×200 magnification. Scale bar = 200 μm.

**Supplemental Figure 4. Tissue damage in scraped centers at Tissue Select settings 0–4.**
Representative images at ×200 magnification show the central regions of the CUSA-scraped areas.
Tissue damage in the adjacent normal ovarian tissue varied by Tissue Select setting:
at setting 0, vacuolization and degeneration extended to 200–400 μm in depth;
at settings 1–2, similar effects were observed but were generally confined within 200 μm;
at settings 3–4, damage was more limited, with minimal changes seen at setting 4. 
Scale bar = 200 μm.

**Supplemental Figure 5. Localization of primordial follicles relative to the endometriotic lesion.**
The left image shows a primordial follicle located approximately 600 μm beneath the endometriotic lesion. The right panel presents a magnified view of the primordial follicle. Scale bar (right image) = 200 μm.

**Supplementary Video Legend**

**Supplemental Video. Demonstration of CUSA scraping applied to the central region of the specimen.** 
The video shows how the CUSA device was used to gently scrape the central area of the endometriotic cyst wall. A visible color change occurs in the treated tissue, which may serve as a practical intraoperative indicator that scraping has been performed.
