## Supplementary_Methods for "Selective Removal of Endometriotic Lesions Using CUSA Clarity in Ovarian Endometriomas: A Case-Based Histopathological Study"

**Materials & Methods**

Immunohistochemical staining

Paraffin embedded specimens were sectioned at 4 μm thickness and mounted on glass slides. After deparaffinization and rehydration, antigen retrieval was performed using 10 mM Tris / 1 mM EDTA buffer (pH 9.0) at 121 °C for 20 minutes. Endogenous peroxidase activity was blocked using 3% hydrogen peroxide for 10 minutes.
For immunohistochemical staining, slides were incubated overnight at 4 °C with the following primary antibodies:
– anti-cytokeratin 7 (CK7) rabbit monoclonal antibody (1:4000, clone EPR17078, ab181598, Abcam, UK)
– anti-CD10 mouse monoclonal antibody (1:200, clone 56C6, CD10/CALLA [Neutral Endopeptidase] ab-2, MS-728-50, Thermo Fisher Scientific, USA)
Immunoreactivity was detected using the avidin–biotin–peroxidase complex method according to the manufacturer’s instructions (VECTASTAIN ABC Kit, Vector Laboratories, USA). Diaminobenzidine (DAB) was used as the chromogen, and hematoxylin was used for counterstaining.

CUSA application

Tissue removal was performed using the CUSA® Clarity system (Integra LifeSciences, USA) equipped with a 23 kHz handpiece. The central region of each specimen was gently scraped under direct visualization. Five different Tissue Select® settings (0–4) were evaluated, with one specimen section assigned to each setting.
The amplitude was fixed at 65%, aspiration at 40%, and irrigation rate at 3 mL/min. Only the Tissue Select® setting was varied across samples, while all other parameters remained constant throughout the procedure.
